# Morphometric variations of human mandible in Indian population: comparison between subjects having healthy and ankylosed temporomandibular joint

**DOI:** 10.1101/2024.09.23.24314231

**Authors:** Girish Chandra, Rajdeep Ghosh, Kamalpreet Kaur, Ajoy Roychoudhury, Sudipto Mukherjee, Anoop Chawla, Kaushik Mukherjee

**Affiliations:** Department of Mechanical Engineering, Indian Institute of Technology Delhi, New Delhi 110 016, Delhi, India; Department of Mechanical Engineering, The University of Sheffield, Sheffield S1 3JD, United Kingdom; INSIGNEO Institute for in silico Medicine, The University of Sheffield, Sheffield S1 3JD, United Kingdom; Department of Oral & Maxillofacial Surgery CDER, All India Institute of Medical Sciences, New Delhi 110029, Delhi, India

**Keywords:** Morphometry, Temporomandibular joint, Ankylosis, Mandibular implants, Fit discrepancy

## Abstract

The temporomandibular joint (TMJ) helps to perform the functional activities of the oral cavity. Such activities often get affected by end-stage degenerative disorders such as TMJ ankylosis. Alloplastic reconstruction using TMJ implants helps to restore those activities. However, commercially available stock implants often suffer from fit discrepancies in Indian population. Therefore, this study is aimed to compare comparison between mandibular morphometry of subjects with healthy or normal TMJ joints (TMJN) and patients with TMJ ankylosis (TMJA) from the Indian population. Furthermore, the observed mandibular morphometry has also been compared with those of the other populations. In this study, the most useful anatomical parameters of mandible are measured from the CT-based reconstructed mandibles of 367 Indian subjects (199 healthy;168 ankylosis). Significant differences in ramus length (healthy males: 61.74±7.53 mm, ankylosis males: 46.81±10.35 mm; healthy females: 55.21±6.12 mm, ankylosis females: 41.77±8.57 mm) and condyle width (healthy males: 18.76±3.22 mm, ankylosis males: 22.67±5.56 mm; healthy females: 16.94±2.41 mm, ankylosis females: 21.31±4.65 mm) have been observed between mandibles of ankylosis and healthy subjects. Differences in ramus length (affected side: 43.87±9.51 mm; unaffected side: 55.34±7.12 mm) and condyle width (affected side: 23±4.68 mm; unaffected side: 17.99±2.81 mm) were also observed between affected and unaffected sides of mandibles for unilateral ankylosis patients. Ramus length in healthy subjects was found to be the most statistically significant parameter between mandibles of Indians and other populations.

## 1. Introduction

Ankylosis of temporomandibular joint (TMJ) is a clinical condition where the condyle of the diseased mandible gets fused with the glenoid fossa. Such a condition interrupts the functional activities related to mouth opening. Genetic etiology, trauma and infection are the most common causes of TMJ ankylosis (TMJA) in children (Durham et al., 2015; Elgazzar et al., 2010; Liu & Steinkeler, 2013; Long et al., 2005; Roychoudhury et al., 2021). Multiple morphological changes (such as shortening of ramus length, debossing of antegonial notch, warping of the mandible, and bulging of condyle) are observed in the jaw of an ankylosis patient due to prolonged restricted jaw mobility and inhibited growth (Arakeri et al., 2012; Kaban et al., 1990). Figure 1 shows such observed changes in case of a representative TMJA patient.

**Figure 1.**
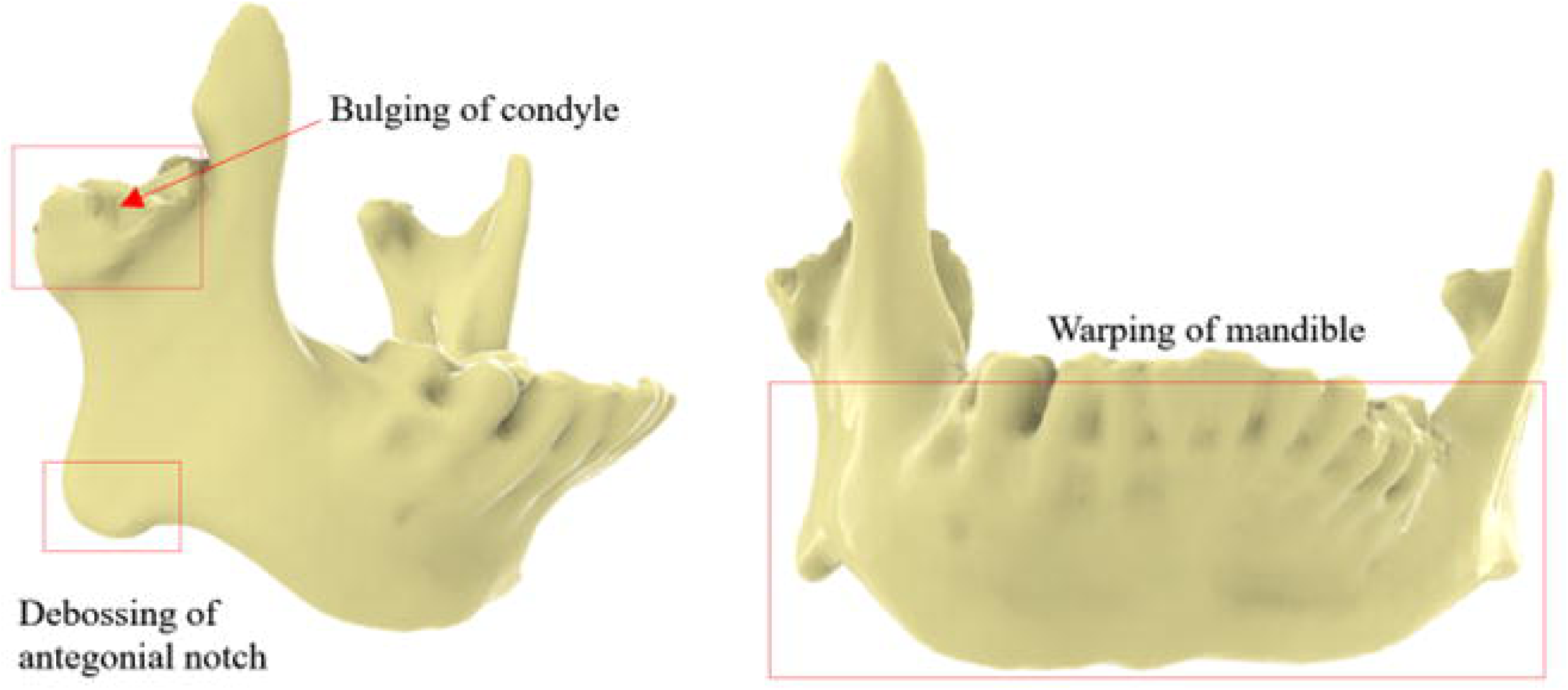
Morphological changes observed in the mandible of an Indian female patient suffering from TMJ ankylosis

Patients suffering from joint degenerative disorders and ankylosis (mostly intermediate and end-stage cases) are often clinically advised for total TMJ replacement surgery (Yoda et al., 2020). Two types of TMJ implants are currently commercially available to restore normal functioning of the joint. The first type is customized patient-specific implants like TMJ Concepts (Stryker Corporation, USA) (Stryker, 2022), while the other one is stock implant like Biomet Microfixation System (Zimmer Biomet Holdings Inc, Jacksonville, FL, USA). To cater the variability in mandibular morphometry, typically these stock implants are available in different shapes and sizes (BIOMET Microfixation, 2007). The Biomet stock prosthesis has 3 fossa baseplate sizes (small, medium, large) and 5 different ramus components (three sizes in standard type: 45mm, 50mm, 55mm; two sizes in narrow type: 45 mm, 50 mm), including an offset ramus variant in all 5 sizes. The stock TMJ implants (especially Biomet TMJ implants) are mostly used in developing and developed countries for TMJA patients due to cost-effectiveness and instant availability (Roychoudhury et al., 2021).

However, recent India-centric study has shown that the commercially available stock TMJ implants exhibit fit discrepancies and require significant bone trimming during surgery (Alagarsamy et al., 2021). This is because most of these stock implants were designed for skeletally mature (mean age of 40 years) Caucasian subjects (Kolte et al., 2023; Walter Lorenz Surgical Incorporated, 2005) whereas the mean age of Indian population suffering from TMJA is much less (24.26 years) (Alagarsamy et al., 2021). Furthermore, only 14% of the patient population considered for designing these stock implants was suffering from ankylosis(Walter Lorenz Surgical Incorporated, 2005). Therefore, the morphological variations due to age and ankylosis might not have been considered in the design of these stock TMJ implants. Although population-specific mandibular morphometric studies have been performed in other countries (Akhlaghi et al., 2014; Vallabh et al., 2020) including India (Sharma et al., 2016), all these studies have considered only the healthy population. To the best of the authors’ knowledge, there is no study investigating the morphometry of mandibles in ankylosis patients and further comparing it with the healthy population.

The primary objective of this present study is to present a comparison between mandibular morphometry of subjects with healthy or normal TMJ (TMJN) and patients with TMJ ankylosis (TMJA) from the Indian population. Furthermore, the observed mandibular morphometry has also been compared with those of the other populations to understand the inter-country anatomical differences.

## 2. Materials and Methods

### 2.1 Dataset and Mandible Segmentation

Clinical CT dataset of randomly selected 199 healthy or control (112 males and 87 females) and 168 TMJ ankylosis (84 males and 84 females) subjects were retrospectively obtained from AIIMS Delhi, India with due ethical approval and informed consent. 3D reconstruction of these dataset was performed in Materialise Mimics v25.0 (Materialise Inc., Leuven, Belgium). 98 ankylosis subjects (51 males and 47 females) had unilateral ankylosis whereas the remaining subjects (33 males and 37 females) had bilateral ankylosis. The complete dataset is thus divided into four groups, namely, TMJ normal/healthy (TMJN) males, TMJN females, TMJ ankylosis (TMJA) males and TMJA females.

### 2.2 Landmarks on mandible and corresponding parameter measurement

Earlier studies on morphometry of human mandibles used several anatomical parameters to determine various dimensions of a healthy human mandible (Dong et al., 2021; Haas et al., 2001; Ongkosuwito et al., 2009). Similar landmarks are also identified in this current study using Materialise 3-Matic (version 17.0) to understand the mandible morphometry in healthy and ankylosis patients. Details of these landmarks are listed in Table 1 and shown in Figure 2.

**Figure 2.**
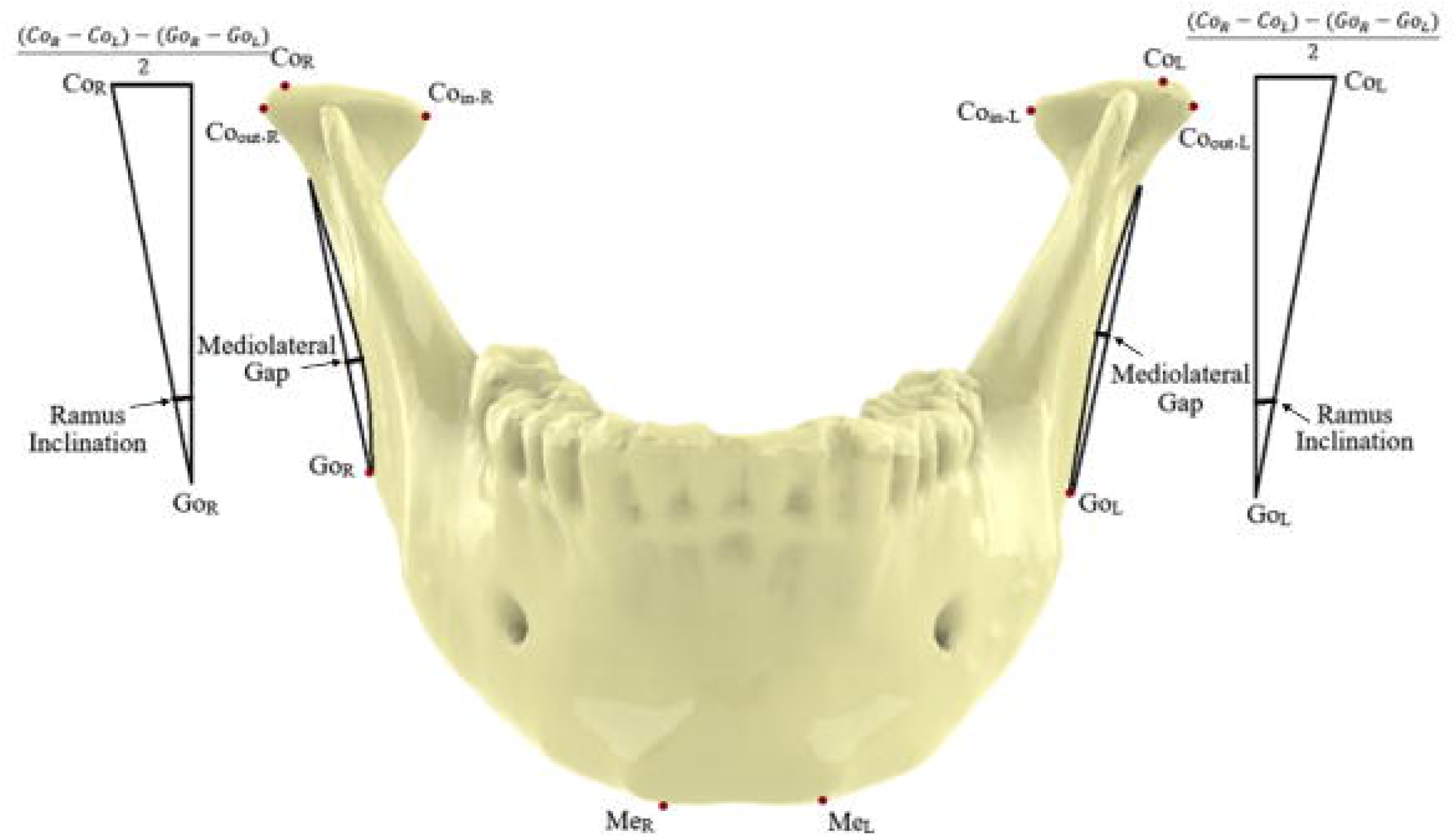
Landmarks chosen on the mandible of a healthy subject

**Table 1.** The landmark used and their definitions on the human mandible for both right (R) and left (L) sides.

| Landmark name | Abbreviation | Definition |
| --- | --- | --- |
| Condylion | Co or Co<sub>R</sub>/Co<sub>L</sub> | The superior-most point on the condylar head |
| Gonion | Go or Go<sub>R</sub>/Go<sub>L</sub> | The most postero-inferior point on mandible angle |
| Menton | Me or Me<sub>R</sub>/Me<sub>L</sub> | The inferior-most point on mandible symphysis |
| Lateral Condyle | Co<sub>outR</sub>/ Co<sub>outL</sub> | The lateral-most point on the condylar head |
| Medial Condyle | Co<sub>inR</sub> / Co<sub>inL</sub> | The medial-most point on the condylar head |

Ramus length, minimum ramus width, gonion angle, condyle width, intergonial distance, intercondyle distance, ramus inclination, and mediolateral gap (measure of anatomical curvature at the ramus) were considered important morphometric parameters of mandible (Ackland et al., 2017; Vallabh et al., 2020). These parameters and their methods of measurement are listed in Table 2. In ankylosed sides of TMJA patients, the joints being fused and severely damaged, there was no distinct landmark to measure intercondyle distance and to calculate ramus inclination, and mediolateral gap. Therefore, the measurements of these morphometric parameters were excluded for TMJA subjects.

**Table 2.** Anatomical parameters and their measuring methods in the human mandible.

| Measured Parameters | Notation | Measuring Methods |
| --- | --- | --- |
| Ramus Length | Co-Go | Euclidean distance between the condylion and gonion for both sides |
| Minimum Ramus Width | MRW | Minimum Euclidean distance between the anterior and posterior points in the ramus for both sides |
| Gonion Angle (GA) | $\angle \text{CoGoMe}$ | The angle between condylion, gonion and menton |
| Condyle width (CW) | $\text{Co}_{\text{in}} - \text{Co}_{\text{out}}$ | Euclidean distance between medial and lateral condyles. |
| Intergonial Distance (IGD) | $\text{Go}_R - \text{Go}_L$ | Euclidean distance between left and right gonions. |
| Intercondyle Distance (ICD) | $\text{Co}_R - \text{Co}_L$ | Euclidean distance between left and right condylion |
| Ramus inclination | RI | $\text{RI} = \sin^{-1} \left( \frac{\frac{(\text{Co}_R - \text{Co}_L) - (\text{Go}_R - \text{Go}_L)}{2}}{\text{Co} - \text{Go}} \right)$ |
| Mediolateral Gap | MG | $\text{MG} = \frac{\text{arc length of mediolateral curve}}{\varnothing} (1 - \cos \frac{\varnothing}{2})$<br>where $\varnothing = \text{arc angle}$ and $\frac{\sin(\frac{\varnothing}{2})}{\varnothing} = \frac{\text{chord length of mediolateral curve}}{2 \times \text{arc length of mediolateral curve}}$ |

To minimize systematic errors in measurements, an inter-operator variability study was also performed by two operators for ramus length. A Bland-Altman plot for measurement of ramus length is shown in Figure 3. A significant coefficient of determination (R^2^=0.9988) has been observed between the measurements done by two independent operators. Inter-operator difference in measurements was observed to have a mean absolute error of 0.06 mm (0.1 % of mean ramus length) and standard deviation of 0.25 mm (0.4 % of mean ramus length). This establishes confidence in the correctness of the dimensions reported in the present study.

**Figure 3.**
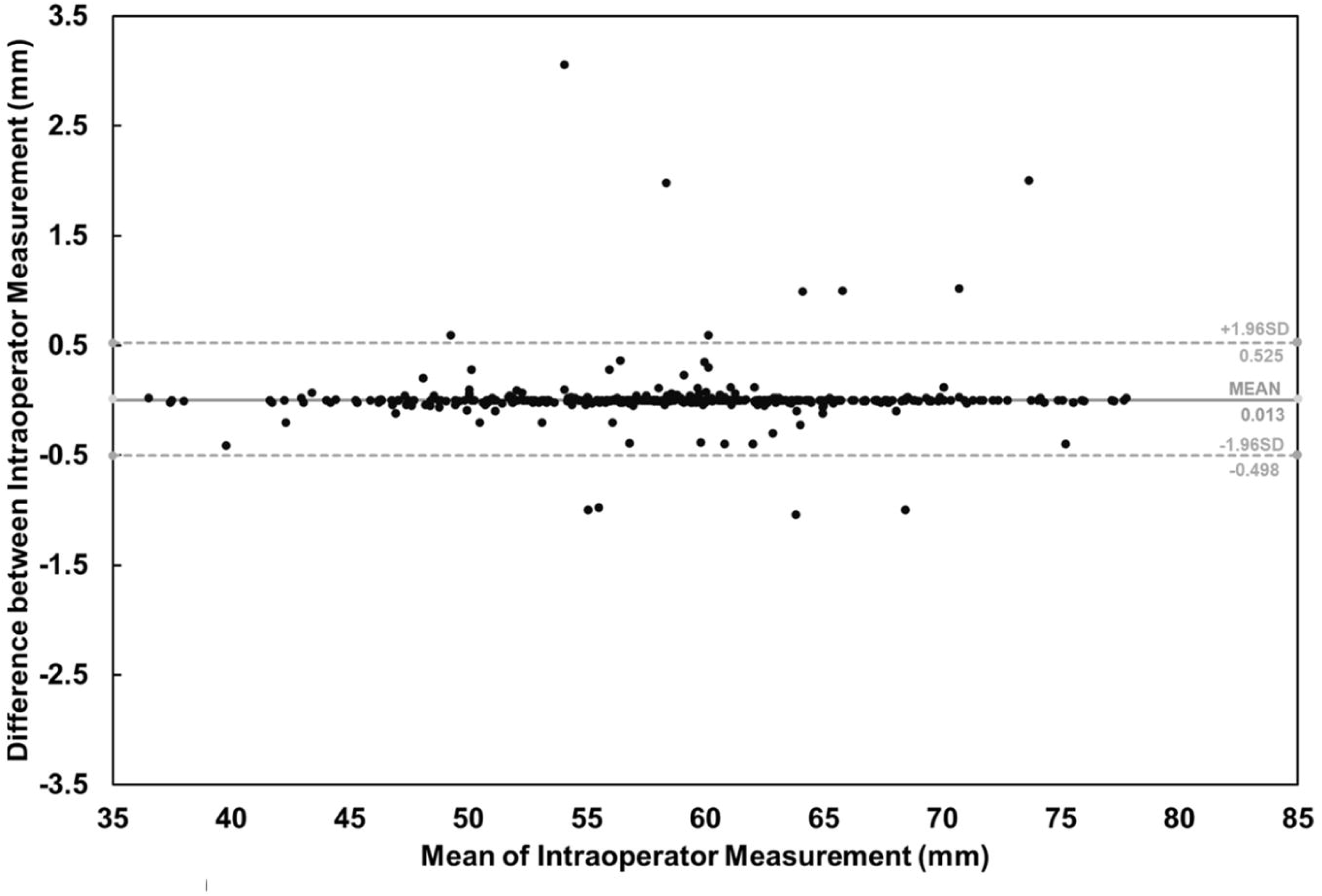
Bland-Altman plot with its limits of agreement for ramus length

### 2.3 Statistics, comparisons, and their hypotheses

To obtain the minimum sample size sufficient to represent a population, the Cochran formula (Equation 1) was used with an estimated proportion of the population (*p*) suffering from TMJ ankylosis(Uakarn et al., 2021). A previous epidemiological study on 21,720 children in Lucknow, India reported that 0.046% of them suffered from TMJ ankylosis (Gupta et al., 2012). Due to unavailability of information regarding percentage of Indian population suffering from TMJ ankylosis, a maximum of 1% Indian population is assumed to be suffering from TMJ ankylosis (p). With a 2% error margin and 90% confidence interval, a sample size of 53 were found sufficient using Cochran’s method (eq. 1).

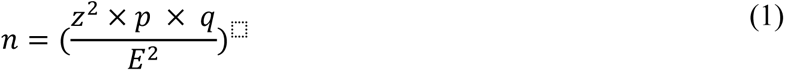

Where *p* is the estimated proportion of population suffering from TMJ ankylosis, *q* is the estimated proportion of population not suffering from TMJ ankylosis (healthy subjects), *z* value is based on a confidence level, and *E* is the allowable margin of error.

The measured parameters for the different groups of Indian population (TMJN male, TMJN female, TMJA male, and TMJA female) have been compared with one another as well as with morphology reported in earlier literature containing data from other countries (Choi et al., 2011; Dong et al., 2021; Lopez-Capp et al., 2018; Moskowitch et al., 1993; Vallabh et al., 2020). For the chosen morphometric parameters, the following hypotheses were tested using two-sample z-test (95% significant interval) based on mean and known variance of the sample sizes from two different groups of subjects (Montgomery et al., 2016):

1. There is no statistically significant difference in mandibular morphometry between healthy and ankylosis subjects of all age cohorts for both

a. Indian male and
b. Indian female.
2. There is no statistically significant difference in mandibular morphometry between affected and unaffected sides of Indian unilateral ankylosis patients.
3. There is no statistically significant difference in mandibular morphometry between healthy subjects of India and other countries for both (adults)

a. Male and
b. Female groups.

## 3. Results

### 3.1 Morphometry of human mandibles among Indian population

Anthropometric measurements of different mandibular parameters were found to vary with gender (Mehta et al., 2020) as well as age (Rachmadiani et al., 2017). Previous literature reported the effect of age on skeletal maturity (Saggese et al., 2002; Satoh & Hasegawa, 2022). Puberty plays a key role in the development of bones and skeletal mass of an individual (Saggese et al., 2002). Therefore, data of Indian population (TMJN male, TMJN female, TMJA male, and TMJA female) were further subdivided based on age (pre-puberty: <12 years; puberty: 12-18 years; adult: >18 years) to study age-based variations among them.

#### 3.1.1 Influence of age

The extremum (min/max), mean and standard deviations (SD) of the morphometric parameters of the different groups according to three age cohorts are listed in Table 3. In addition, Figure 4 shows the comparison of anthropometric measurements of different mandible parameters of healthy subjects of same gender between ankylosis and healthy subjects in all age cohorts of the Indian population. The mean ramus length increases with age in each group. The mean minimum ramus width and intergonial distance also increase with age in each group. The mean condyle width increases with age in each group, except for TMJA females. The mean intercondyle distance increases with age in both TMJN males and females. The mean mediolateral gap also increases with age in both TMJN males and females. On the contrary, the mean gonion angle decreases with age in each group, except for TMJA males. Negligible change was observed in ramus inclination with age for both TMJN males and females.

**Table 3.**
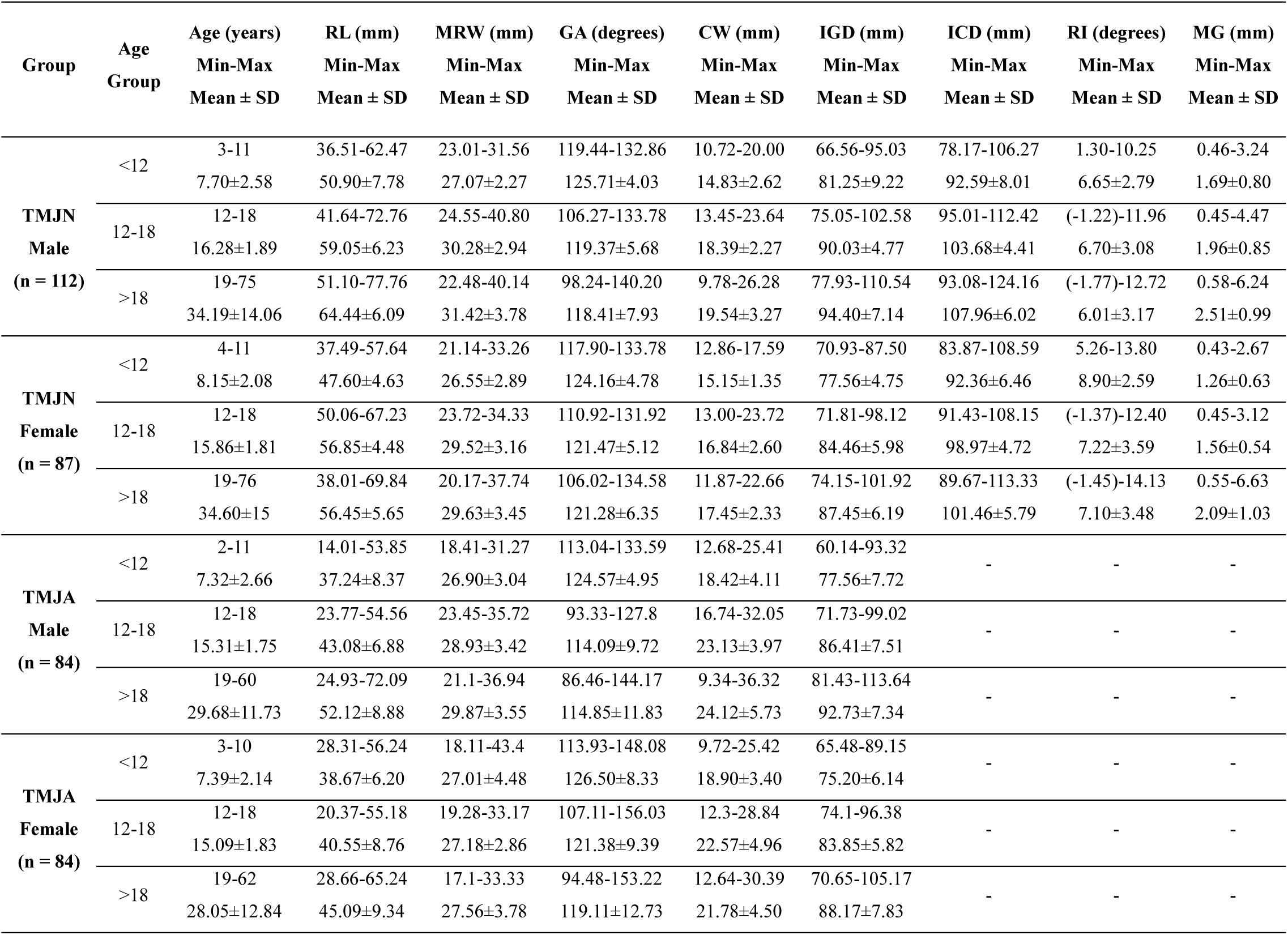
Indian mandible morphometry according to age groups based on parameters of 199 TMJN and ankylosed sides (70 Bilateral & 98 Unilateral) of 168 TMJA subjects.

| Group | Age Group | Age (years) | RL (mm) | MRW (mm) | GA (degrees) | CW (mm) | IGD (mm) | ICD (mm) | RI (degrees) | MG (mm) |
| --- | --- | --- | --- | --- | --- | --- | --- | --- | --- | --- |
|  |  | Min-Max | Min-Max | Min-Max | Min-Max | Min-Max | Min-Max | Min-Max | Min-Max | Min-Max |
| | | Mean ± SD | Mean ± SD | Mean ± SD | Mean ± SD | Mean ± SD | Mean ± SD | Mean ± SD | Mean ± SD | Mean ± SD |
| **TMJN Male**<br>(n = 112) | <12 | 3-11 | 36.51-62.47 | 23.01-31.56 | 119.44-132.86 | 10.72-20.00 | 66.56-95.03 | 78.17-106.27 | 1.30-10.25 | 0.46-3.24 |
| | | 7.70±2.58 | 50.90±7.78 | 27.07±2.27 | 125.71±4.03 | 14.83±2.62 | 81.25±9.22 | 92.59±8.01 | 6.65±2.79 | 1.69±0.80 |
|  | 12-18 | 12-18 | 41.64-72.76 | 24.55-40.80 | 106.27-133.78 | 13.45-23.64 | 75.05-102.58 | 95.01-112.42 | (-1.22)-11.96 | 0.45-4.47 |
| | | 16.28±1.89 | 59.05±6.23 | 30.28±2.94 | 119.37±5.68 | 18.39±2.27 | 90.03±4.77 | 103.68±4.41 | 6.70±3.08 | 1.96±0.85 |
|  | >18 | 19-75 | 51.10-77.76 | 22.48-40.14 | 98.24-140.20 | 9.78-26.28 | 77.93-110.54 | 93.08-124.16 | (-1.77)-12.72 | 0.58-6.24 |
| | | 34.19±14.06 | 64.44±6.09 | 31.42±3.78 | 118.41±7.93 | 19.54±3.27 | 94.40±7.14 | 107.96±6.02 | 6.01±3.17 | 2.51±0.99 |
| **TMJN Female**<br>(n = 87) | <12 | 4-11 | 37.49-57.64 | 21.14-33.26 | 117.90-133.78 | 12.86-17.59 | 70.93-87.50 | 83.87-108.59 | 5.26-13.80 | 0.43-2.67 |
| | | 8.15±2.08 | 47.60±4.63 | 26.55±2.89 | 124.16±4.78 | 15.15±1.35 | 77.56±4.75 | 92.36±6.46 | 8.90±2.59 | 1.26±0.63 |
|  | 12-18 | 12-18 | 50.06-67.23 | 23.72-34.33 | 110.92-131.92 | 13.00-23.72 | 71.81-98.12 | 91.43-108.15 | (-1.37)-12.40 | 0.45-3.12 |
| | | 15.86±1.81 | 56.85±4.48 | 29.52±3.16 | 121.47±5.12 | 16.84±2.60 | 84.46±5.98 | 98.97±4.72 | 7.22±3.59 | 1.56±0.54 |
|  | >18 | 19-76 | 38.01-69.84 | 20.17-37.74 | 106.02-134.58 | 11.87-22.66 | 74.15-101.92 | 89.67-113.33 | (-1.45)-14.13 | 0.55-6.63 |
| | | 34.60±15 | 56.45±5.65 | 29.63±3.45 | 121.28±6.35 | 17.45±2.33 | 87.45±6.19 | 101.46±5.79 | 7.10±3.48 | 2.09±1.03 |
| **TMJA Male**<br>(n = 84) | <12 | 2-11 | 14.01-53.85 | 18.41-31.27 | 113.04-133.59 | 12.68-25.41 | 60.14-93.32 | - | - | - |
| | | 7.32±2.66 | 37.24±8.37 | 26.90±3.04 | 124.57±4.95 | 18.42±4.11 | 77.56±7.72 | - | - | - |
|  | 12-18 | 12-18 | 23.77-54.56 | 23.45-35.72 | 93.33-127.8 | 16.74-32.05 | 71.73-99.02 | - | - | - |
| | | 15.31±1.75 | 43.08±6.88 | 28.93±3.42 | 114.09±9.72 | 23.13±3.97 | 86.41±7.51 | - | - | - |
|  | >18 | 19-60 | 24.93-72.09 | 21.1-36.94 | 86.46-144.17 | 9.34-36.32 | 81.43-113.64 | - | - | - |
| | | 29.68±11.73 | 52.12±8.88 | 29.87±3.55 | 114.85±11.83 | 24.12±5.73 | 92.73±7.34 | - | - | - |
| **TMJA Female**<br>(n = 84) | <12 | 3-10 | 28.31-56.24 | 18.11-43.4 | 113.93-148.08 | 9.72-25.42 | 65.48-89.15 | - | - | - |
| | | 7.39±2.14 | 38.67±6.20 | 27.01±4.48 | 126.50±8.33 | 18.90±3.40 | 75.20±6.14 | - | - | - |
|  | 12-18 | 12-18 | 20.37-55.18 | 19.28-33.17 | 107.11-156.03 | 12.3-28.84 | 74.1-96.38 | - | - | - |
| | | 15.09±1.83 | 40.55±8.76 | 27.18±2.86 | 121.38±9.39 | 22.57±4.96 | 83.85±5.82 | - | - | - |
|  | >18 | 19-62 | 28.66-65.24 | 17.1-33.33 | 94.48-153.22 | 12.64-30.39 | 70.65-105.17 | - | - | - |
| | | 28.05±12.84 | 45.09±9.34 | 27.56±3.78 | 119.11±12.73 | 21.78±4.50 | 88.17±7.83 | - | - | - |

**Figure 4.**
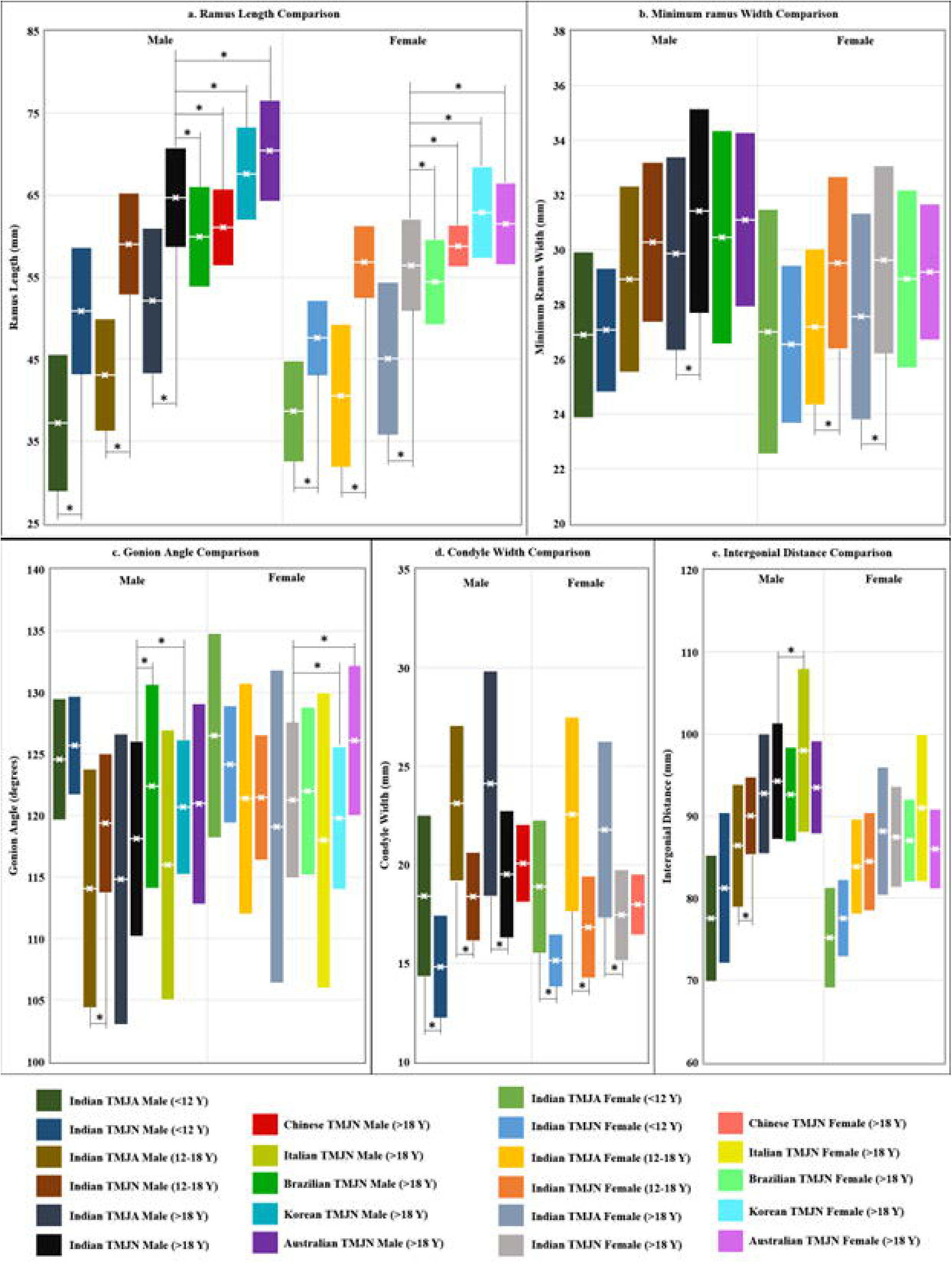
Comparison between Indian population (healthy and ankylosis of all age cohorts) and population (adult) from other countries for (a) ramus length, (b) minimum ramus width, (c) gonion angle, (d) condyle width, and (e) intergonial distance. (* indicates the statistically significant difference with a 95% significance level)

#### 3.1.2 Influence of gender

The following section describes the comparison of morphometric parameters of mandible considering all age cohorts. The mean ramus length was higher in males (TMJN: 61.74±7.53 mm, TMJA: 46.81±10.35 mm,) than in females (TMJN: 55.21±6.12 mm, TMJA: 41.77±8.57 mm). TMJA group exhibited reduced ramus length as compared to TMJN. The mean minimum ramus width is observed to be higher in males (TMJN: 30.67±3.63 mm, TMJA: 29±3.59 mm) than in females (TMJN: 29.14±3.46 mm, TMJA: 27.21±3.73 mm). Ankylosis was also found to reduce minimum ramus width irrespective of gender. While a marginally higher gonion angle was observed in females (TMJN: 121.76±5.90°, TMJA: 121.99±10.78°) as compared to males (TMJN: 119.34±7.35°, TMJA: 116.69±10.95°), gonion angle was found not to be influenced by ankylosis. Mean condyle width (Table 3) was also observed to be higher for males (TMJN: 18.76±3.22 mm, TMJA: 22.67±5.56 mm) than females (TMJN: 16.94±2.41 mm, TMJA: 21.31±4.65 mm). In addition, condyle width was also observed to be higher in the ankylose subjects than healthy ones for both genders. Mean intergonial distance was observed to be higher in males (TMJN: 91.89±7.72 mm, TMJA: 87.78±9.42 mm) than females (TMJN: 85.14±6.84 mm, TMJA: 82.94±8.35 mm). Additionally, a reduction in mean intergonial distance was observed due to ankylosis. Mean intercondyle distance and mediolateral gap were observed to be higher in males (intercondyle distance: 105.33±7.23 mm, mediolateral gap: 2.27±0.97 mm) than females (intercondyle distance: 99.43±6.43 mm, mediolateral gap: 1.84±0.93 mm) of healthy group (TMJN), while mean ramus inclination was observed to be lower in males (TMJN: 6.30±3.13°) than females (TMJN: 7.41±3.43 mm).

#### 3.1.3 Distribution of morphometric parameters

Figure 5 shows the distribution of each parameter (for both TMJN and TMJA groups) considering all age cohorts. Number of TMJA subjects with ramus length less than 45 mm and condyle width more than 18 mm were significantly higher than those of TMJN subjects (Figure 4(a), (d)). However, no such significant difference between TMJN and TMJA groups was observed for minimum ramus width and gonion angle (Figure 5 (b) and (c)). Furthermore, distribution of ramus inclination and mediolateral gap are shown in Figure 5 (e) and (f) respectively for the healthy subjects only as it was not possible to measure these parameters in the ankylosis subjects due to severe warping of mandibles.

**Figure 5.**
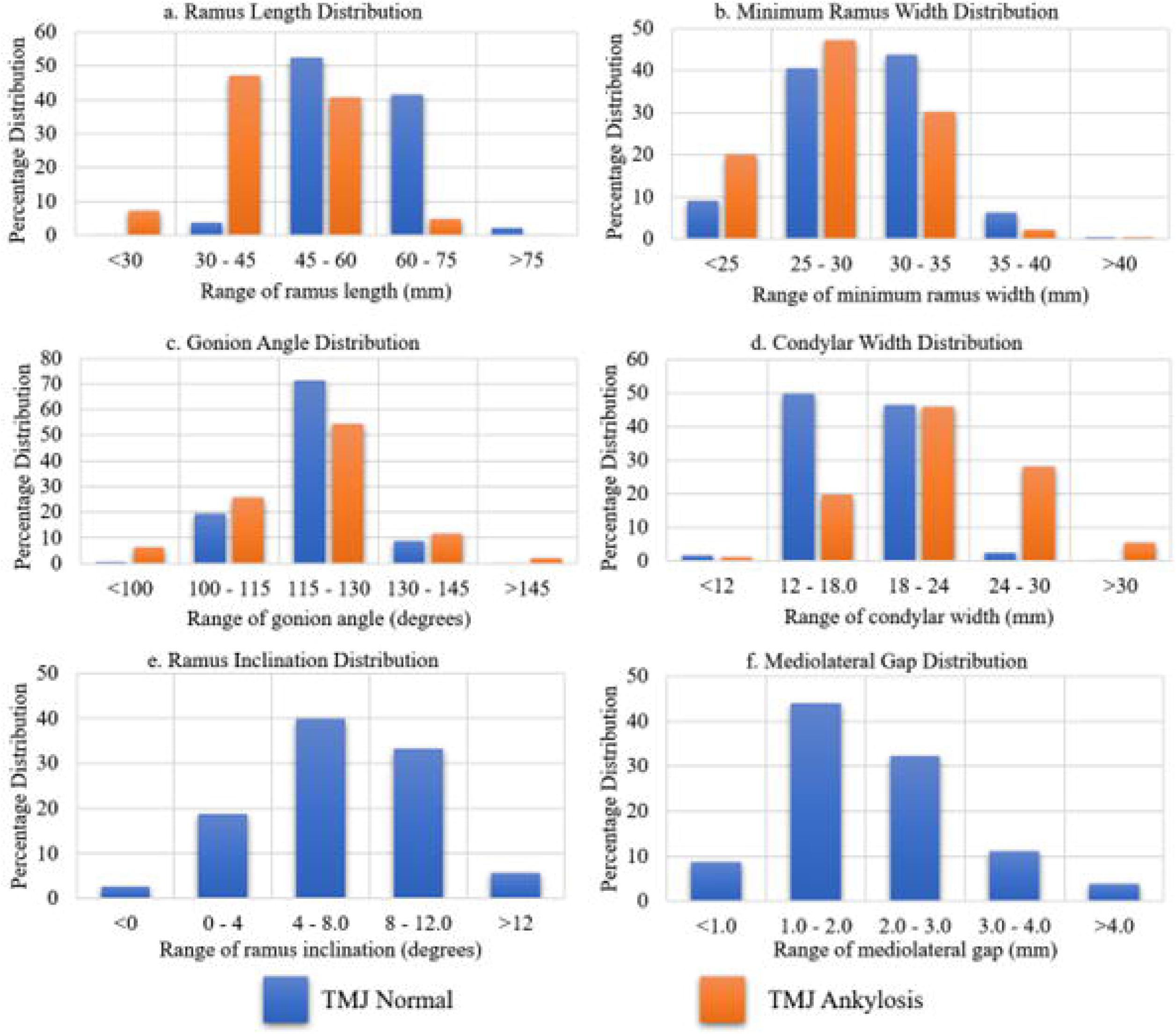
Variation of morphometric parameters in TMJN and TMJA patients (a) ramus length, (b) minimum ramus width, (c) gonion angle, (d) condyle width, (e) ramus inclination, and (f) mediolateral gap

### 3.2 Hypothesis 1: There is no statistically significant difference in morphometry of mandible between healthy and ankylosis subjects of Indian population

A gender- and age-matched comparison of mandible morphometry between ankylosis and healthy subjects of Indian population from all age cohort is presented in Figure 4. Our analysis indicates significant statistical differences (*) in ramus length (male and female of all age cohorts), minimum ramus width (male for adult & female for puberty and adult), gonion angle (male for puberty only), condyle width (male and female of all age cohorts), and intergonial distance (male of puberty only) between healthy and ankylosis subjects of Indian population (Figure 4). For other morphometric parameters like minimum ramus width (male for pre-puberty and puberty & female for pre-puberty), gonion angle (male for pre-puberty and adult & female for all age cohorts) and intergonial distance (male for pre-puberty and adult & female for all age cohorts), no statistically significant difference was observed. Based on this analysis, it was established that mandibular morphometry in subjects with ankylosis differs from those of the healthy population in India and, therefore, hypothesis 1 was rejected.

### 3.3 Hypothesis 2: There is no statistically significant difference in mandibular morphometry between affected and unaffected sides of Indian unilateral ankylosis patients

To test hypothesis 2, all the morphometric parameters were compared between affected and unaffected sides of unilateral ankylosis patients. The affected side exhibited a reduced ramus length (affected side: 43.87±9.51 mm; unaffected side: 55.34±7.12 mm), reduced gonion angle (affected side: 116.89±9.81°; unaffected side: 119.58±5.72°) and an increased condyle width (affected side: 23±4.68 mm; unaffected side: 17.99±2.81 mm) as compared to the unaffected side in such unilateral ankylosis cases. No such difference in minimum ramus width was, however, observed between the affected and unaffected sides. Since the differences were statistically significant, hypothesis 2 was rejected and it was established that ankylosis influences the mandibular morphometry of the affected side.

### 3.4 Hypothesis 3: There is no statistically significant difference in mandible morphometry between the healthy population of India and other countries

A gender- and age-matched comparison of mandible morphometry between healthy adult population of India and other countries like Brazil (Lopez-Capp et al., 2018), China (Dong et al., 2021), Korea (Choi et al., 2011), Australia (Vallabh et al., 2020), and Italy (Moskowitch et al., 1993) is also presented in Figure 4. The statistical test exhibited that ramus length of Indian healthy population is significantly different than those of all other countries (Figure 4-a). The gonion angle in Indian healthy population was found to be statistically different than populations of Brazil (male), Korea (male and female) and Australia (female). The intergonial distance was also found to be statistically different than those of Italy (male). No statistical difference was observed in other mandibular morphometric parameters in healthy populations of India and other countries. Thus, it was established that the mandible morphometry of Indian population differed from other countries like Brazil, China, Korea, Australia and Italy, and therefore, hypothesis 3 was rejected.

## 4. Discussion

The primary goal of this study was to investigate the difference in morphometric parameters of human mandibles between healthy and TMJ ankylosed Indian subjects. In addition, the present study also reports the differences in mandibular morphometry between population of India and other countries The results of this investigation might provide key insights on the possible design modifications in commercially available TMJ implants focusing on Indian patients suffering from TMJ ankylosis.

Higher mean ramus length was observed in Indian males as compared to those in females. Similar observations were also reported in previous studies on populations from India (Mehta et al., 2020) and other countries (Choi et al., 2011; Dong et al., 2021; Lopez-Capp et al., 2018; Vallabh et al., 2020). Earlier studies (Markande et al., 2012; Rai et al., 2007) also reported that ramus length exhibits a higher sexual dimorphism than any other morphometric parameters of the human mandible. However, mean RL in Indian TMJA population was found to be smaller (up to a maximum of 40% with mean values) than Indian healthy population. It should be noted that the minimum ramus lengths in pre-puberty, puberty and adult were 14.01 mm, 20.37 mm, and 24.93 mm, respectively. These dimensions are way smaller than the minimum length (45 mm) of the existing mandibular component of Biomet TMJ implants (BIOMET Microfixation, 2007). In addition, the present study also reports a significant portion (more than 50%) of Indian population suffering from TMJA whose ramus length was less than 45mm.

Like ramus length, the minimum ramus width was observed to be higher in Indian males than females. This observation was also consistent with the findings of earlier studies on Indian subjects (Gurushanthappa & Rajashekarappa, 2013; Sharma et al., 2016) as well as subjects from other countries (Lopez-Capp et al., 2018; Vallabh et al., 2020). Furthermore, MRW in Indian TMJA was also found to be narrower (up to a maximum of 13.4 % with mean values) than those in Indian TMJN population.

The gonion angle is observed to be higher in females than males similar to the earlier study on Indian population (Mehta et al., 2020) as well as population from other countries (Choi et al., 2011; Lopez-Capp et al., 2018; Moskowitch et al., 1993; Vallabh et al., 2020). Furthermore, similar to the study by Rai et al. (2007), Indian subjects were found to have lesser gonion angle than those of the Caucasians. The present study reported a mean condyle width ranging from 17.45 mm to 19.45 mm in adults which is consistent with the data (16.90 mm to 19.74 mm) obtained from a recent pilot study by Kolte et al. (2023) on retrospective cohorts of subjects who visited a clinical setup in the Indian state of Maharashtra. Although studies are scarce on the measurement of condyle width in Indian subjects, the results from this recent study (Kolte et al., 2023) provide confidence in our results. The condyle width measured in the Indian ankylosis group was also found to be wider (up to a maximum of 29.9 % with mean values) than in the healthy cohort. On the contrary, the intergonial distance measured in the Indian TMJA group was also found to be narrower (up to a maximum of 13.6 % with mean values) than in the TMJN group. Apart from directly measurable mandible parameters, the present study also reported ramus inclination (more than 50% with less than 3 mm) and mediolateral gap (more than 50% with higher than 4°) in Indian TMJN populations. Both these parameters may also be very important from the perspective of the design of stock TMJ implants as also observed before (Alagarsamy et al., 2021).

However, this study still has certain limitations. Although enough care has been taken to minimize bias in the data procured, the obtained data was from a particular clinical center. In future, a multi-centric study may be planned and executed. Precise dimensioning of anatomical parameters highly relies on the experience of the operators and their measuring techniques (Coombs et al., 2019) in taking the measurements. High variance in the measured parameters caused by inexperienced operators may also lead to erroneous interpretations of the results obtained in the study. Nonetheless, the results of this investigation might be useful in determining the necessity of any design modifications in commercially available TMJ implants for Indian patients suffering from TMJ ankylosis. Additionally, such a study might assist anthropologists, forensic investigators, anatomists, and maxillofacial surgeons in providing anthropological and clinical data that might be further useful in medico-legal and dental procedures (Ishwarkumar et al., 2017).

## 5. Conclusion

This work presents a comparative mandible morphometry study on healthy and ankylosis Indian subjects where major anatomical parameters and associated landmarks were chosen to study the anthropometry of mandibles. A statistically significant difference was observed in the measured ramus length and condyle width between the healthy and ankylosis subjects, and between the affected and unaffected sides in unilateral ankylosis subjects. When compared with the morphometry of other populations reported in erstwhile studies, a statistically significant difference was observed mostly in ramus length and gonion angle. Unusual variations of ramus length between healthy and ankylosis subjects along with ramus inclination and mediolateral gap indicate the need to develop stock TMJ implants for the Indian population suffering from TMJ ankylosis.

## Ethical Approval

Required ethical approval was already in place from the Institute Ethics Committee (IEC) of AIIMS, New Delhi, India.

## Acknowledgment and Funding

The authors are thankful to IIT Delhi and AIIMS Delhi for the facilities extended, and the Indo-German Science and Technology Centre (Grant no. IGSTC/Call 2020/add-bite/48/2021-22/260) for the funding.

## Author Contributions

**Girish Chandra:** Conceptualization, Data Curation, Formal Analysis, Investigation, Methodology, Software, Visualization, Writing – original draft. **Rajdeep Ghosh:** Conceptualization, Investigation, Methodology, Software, Validation, Writing – review & editing. **Kamalpreet Kaur:** Resources, Writing – review & editing. **Ajoy Roychoudhury:** Funding acquisition, Project Administration, Resources, Supervision. **Sudipto Mukherjee:** Conceptualization, Funding acquisition, Methodology, Project Administration, Resources, Supervision, Writing – review & editing. **Anoop Chawla:** Conceptualization, Funding acquisition, Methodology, Project Administration, Resources, Supervision, Writing – review & editing. **Kaushik Mukherjee:** Conceptualization, Funding acquisition, Methodology, Project Administration, Resources, Supervision, Writing – review & editing.

## Data Availability Statement

The data that support the findings of this study are available from the corresponding author upon reasonable request.

